# Laboratory evidence of retained immunity to exanthematous viruses in primary and secondary immunodeficiency

**DOI:** 10.1101/2025.03.13.25323766

**Authors:** Kyriakos Ioannou, Emmanouil Karofylakis, Salma Alkhammash, Helen Baxendale, Sarah Eisen, Rainer Doffinger, Anita Chandra, Ania Manson, James Thaventhiran, Effrossyni Gkrania-Klotsas, Dinakantha Kumararatne

## Abstract

**Background:** Waning immunity to common exanthematous viruses has been observed in vaccinated immunocompetent persons and adult-onset secondary immunodeficiency patients. However, there is a paucity of data on humoral immunity from adult-onset primary immunodeficiency patients.

**Objective:** Assessment of antibody seroprevalence to common exanthematous viruses in adult-onset common variable immunodeficiency (CVID) and secondary antibody deficiency (SeAD).

**Methods:** Retrospective evaluation of IgG levels against tetanus, measles, mumps, rubella and varicella zoster virus, and pre/postvaccination IgG levels against *Haemophilus influenzae* type b (Hib) and a pneumococcal serotype panel.

**Results:** Antibody responses from 50 patients with CVID and 49 with SeAD were available. Humoral immunity against exanthematous viruses at diagnosis in the CVID/SeAD cohorts was present in 55.3%/82.2% (measles), 39.6%/73.9% (mumps), 77.8%/93.2% (rubella), 59.6%/82.

2% (varicella zoster). Pneumococcal and Hib postvaccination responses were deficient in the CVID cohort and 28% and 32% in the SeAD cohort.

**Conclusion:** Antibody responses to exanthematous viruses were more commonly retained compared to postvaccination anti-bacterial polysaccharide responses in both cohorts.

**Summary:** Seroprevalence assessment of exanthematous viruses in antibody deficiency patients revealed retained immunity, in contrast to antibacterial postvaccination responses. Retained antiviral immunity was more likely in secondary antibody deficiency patients. These results are suggestive of retained clinical protection against exanthematous viruses.

## Introduction

Common exanthematous viral infections include measles, mumps, rubella and chickenpox. These highly contagious diseases occur mainly in childhood but can also affect immunologically naïve adults, with their severity ranging from mild complications to permanent disability and death (1, 2). While they were once endemic throughout much of the world their incidence now has largely decreased mainly due to the impact of vaccination programs (3, 4). The impact of measles vaccination, specifically, was highlighted in recent findings of declining case-fatality ratios in low- and middle-income countries in the past 30 years (5). However, measles remains a global public health concern, as immunity gaps from inconsistent vaccine coverage and immunodeficiency leave individuals in developed countries (including the UK), susceptible to imported and endogenous outbreaks and can lead to measles resurgence in countries with previously close-to-elimination rates (6, 7).

Protection against an exanthematous viral infection is greatly dependent on neutralising antibody responses (8, 9). Waning antibody titres after measles-mumps-rubella (MMR) vaccination have been repeatedly observed in the general population (10). A variable trend in decreasing anti-MMR IgG seroprevalence has been demonstrated in patients with adult-onset secondary immunodeficiency due to solid organ and hematopoietic cell transplantation (11-13), CAR T-cell immunotherapy (14) and HIV infection (15).

For patients with primary hypogammaglobulinemia, there is a paucity of data on the persistence of anti-exanthematous viral antibodies, casting doubts on their clinical protection level, in contrast to the well-characterized loss of humoral responses against bacterial pathogens leading to recurrent infections and significant morbidity and mortality (16, 17). In fact, there is now a growing evidence base suggesting clinically significant manifestations of MMR infections being more prevalent in this cohort, with the rubella virus, and its attenuated vaccine version, having been frequently identified in granulomas of patients with various types of combined immunodeficiency (18, 19) including CVID (20, 21), while a rubella virus associated episode of fuchs heterochromic iridocyclitis was identified in a fully vaccinated CVID patient (22).

To add to this, the extent of immunity loss to exanthematous viruses compared to bacterial pathogens in this population is not entirely clear at present.

The aim of this study was to assess the persistence of anti-viral (against MMR and VZV) and anti-bacterial antibodies (against *Streptococcus pneumoniae, Haemophilus influenzae* type b and *Clostridium tetani*) in two cohorts of adult-onset CVID and SeAD patients. Our goal was to explore whether protective humoral responses are differentially retained depending on the cause of hypogammaglobulinemia and pathogen type.

## Results

Data from 50 CVID cases (conforming to published case definition (23)) and 49 SeAD cases were available. The identified causes of SeAD included prior rituximab use for various autoimmune indications (n=34), use of non-rituximab immunosuppressants (n=7), CLL (n=3), MGUS (n=2), hematopoietic cell transplantation for haematological malignancy (n=2), and multiple myeloma (n=1). The two cohorts had similar sex distributions (CVID vs SeAD; males; 30/50 vs 32/49). The SeAD cohort was older by approximately a decade at the time of diagnosis (CVID vs SeAD; median age at diagnosis [IQR]; 51.5 [35] years vs 62 [16] years). At baseline the CVID mean IgG level was lower for CVID patients compared to SeAD (2.94 vs 3.07 g/dL). Women had on average a lower mean IgG level compared to men (2.48 vs 3.31 g/dL).

In the CVID cohort, laboratory evidence of immunity against measles, mumps, rubella, VZV, and tetanus was observed in 55.3%, 39.6%, 77.8%, 59.6% and 72.9% respectively. The entire cohort had poor postvaccination responses to pneumococcal and Hib vaccination (Table I). Among the SeAD population, laboratory evidence of immunity against measles, mumps, rubella, VZV and tetanus was observed in 82.2%, 73.9%, 93.2%, 82.2%, and 93.9% respectively. Adequate postvaccination responses to pneumococcal and Hib vaccination were observed in 28% and 32% respectively (Table I). Poor response to the pneumococcal vaccine in this study was defined as achieving 0.35 ug/ml or less level of IgG antibodies against <7/13 pneumococcal serovars, tested, postvaccination. This method of defining vaccine failure has been previously published (24). To add further credence to our observations, Figures 1a, 1b and 1c also show data on the concentration of serovar specific IgG in healthy, CVID and SeAD cohorts against a cut-off of 1.3 ug/ml, the threshold to be achieved against 70% of the serotypes tested, for the response to be categorised as being between the bounds observed in healthy immunised adults, according to US criteria(25). The validity of this criterion has been debated (26, 27). From the data presented, the defective responses in patients with CVID or SeAD, when compared to healthier adult controls is self-evident.

**Table I.** Laboratory evidence of immunity against MMR, VZV, tetanus, and adequate postvaccination responses to pneumococcal and Hib vaccination in a CVID (N_max_=50) and SeAD cohort (N_max_=49) (See the methods sections for criteria of determination of response vs non-response)

|  | CVID cohort (n/N, %)* | SeAD cohort (n/N, %)* |
| --- | --- | --- |
| Measles | 26/47 (55.3) | 37/45 (82.2) |
| Mumps | 19/48 (39.6) | 34/46 (73.9) |
| Rubella | 35/45 (77.8) | 41/44 (93.2) |
| VZV | 28/47 (59.6) | 37/45 (82.2) |
| APA | 0/50 (0) | 14/50 (28) |
| Hib | 0/50 (0) | 16/50 (32) |
| Tetanus | 35/48 (72.9) | 46/49 (93.9) |

APA: anti-pneumococcal antibodies, CVID: common variable immunodeficiency, Hib: *Haemophilus influenzae* type b, MMR: measles, mumps and rubella virus, SeAD: secondary antibody deficiency, VZV: varicella zoster virus,
\* N denotes the total number of patients with available measurements for each variable. Some measurements were not available at baseline and thus N can be equal or less than $N_{\max}$ in each cohort.

**Figure 1a:**
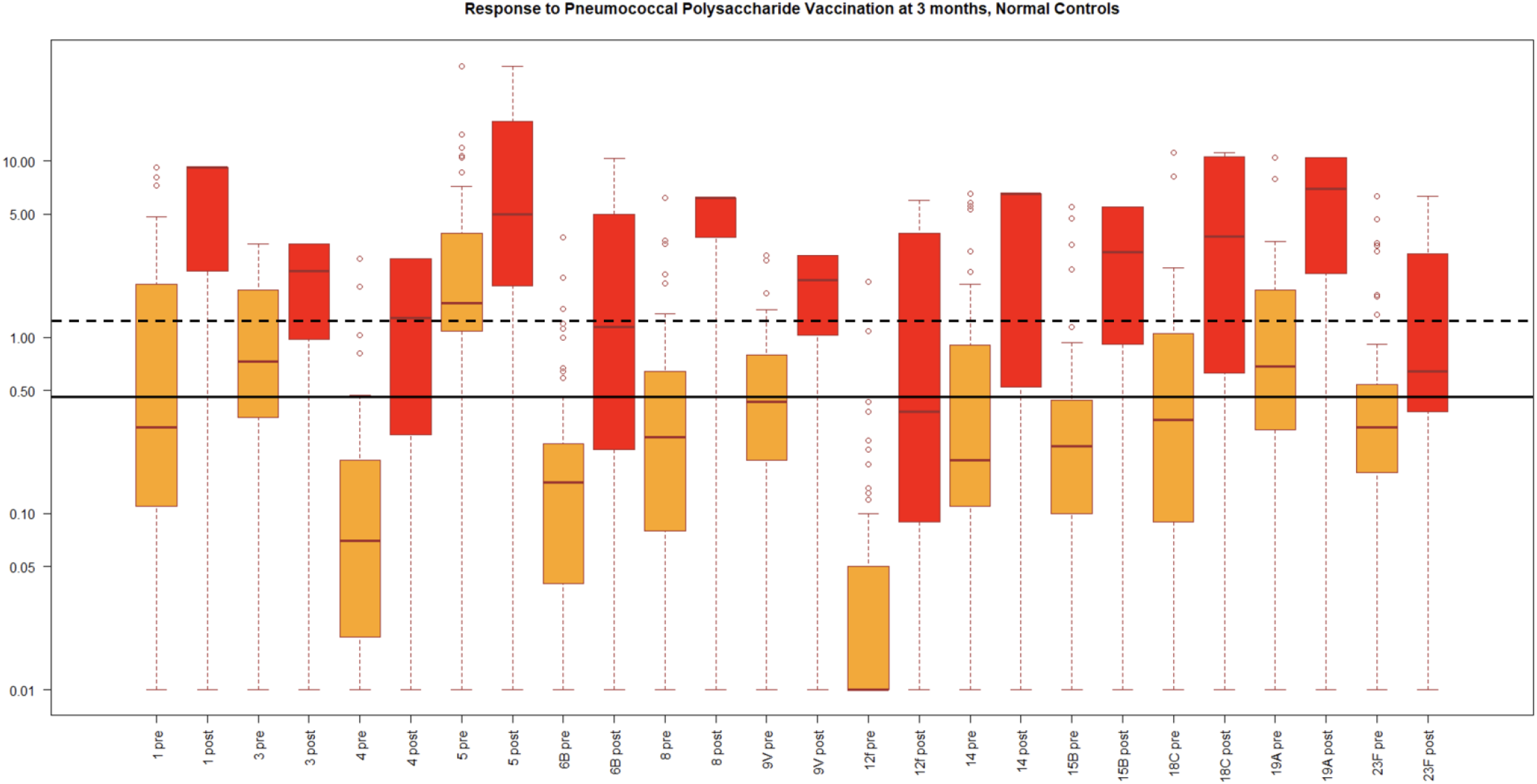
Pneumococcal polysaccharide antibody response of healthy adults: 48 healthy adults aged 18-25 years, who had not previously received pneumococcal vaccine were recruited from the students and staff of University College London, or Great Ormond Street Hospital London and immunised with a single dose of Pneumovax. Serum samples were collected prior to and 3 months after immunisation. Box and Whisker Plots for serotype specific IgG levels across the entire cohort of 48 healthy adults. IgG levels to 13 serotypes (1, 3, 4, 5, 6B, 8, 9V, 14, 18C, 19A, 23F,15B and12F pre- and post-immunization are shown as orange and red bars).The upper and lower quartile for the box and whisker plots are 25% Percentile and 75th percentile, the maximum and minimum observation relates to the 95th and 5th percentiles. The median response for each serotype is indicated by red horizontal line. The putative minimal protective limit of 0.35 ug/ml is indicated by black continuous horizontal line and the limit of 1.3 µg/ml by a black dashed line. (This figure shows unpublished data, cited by courtesy of Dr.H.Baxendale and Dr.Sarah Eisen).

**Figure 1b and 1c:**
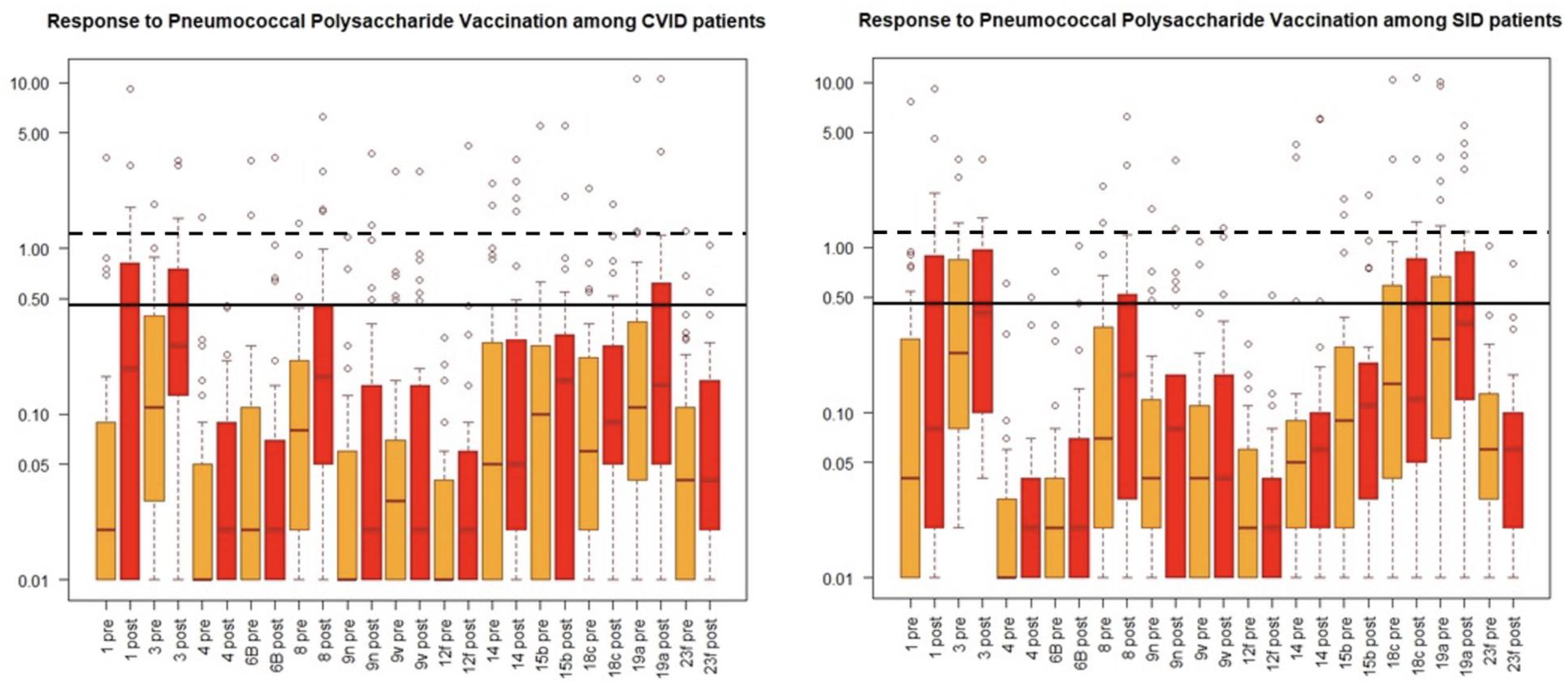
Pneumococcal polysaccharide antibody response of 50 adult CVID patients (1a) and 49 adult SID patients (1b) immunised with a single dose of Pneumovax is shown. Serum samples were collected prior to and 4 weeks after immunisation. Box and Whisker Plots for serotype specific IgG levels across the entire cohort of 48 healthy adults. The median response for each serotype is indicated by red horizontal line. IgG levels to 13 serotypes (1, 3, 4, 6B, 8, 9n, 9V, 14, 15b, 18c, 19a, and 23F) pre- and post-immunisation are shown as orange and red bars. The upper and lower quartile for the box and whisker plots are 25% Percentile and 75th percentile, the maximum and minimum observation relates to the 95th and 5th percentiles. The putative minimal protective limit of 0.35 ug/ml is indicated by black continuous horizontal line and the limit of 1.3 µg/ml by a black dashed line.

After adjusting for age and sex and limiting the analysis among participants with no missing data (n=82), compared to patients with CVID, patients with SeAD were more likely to have retained immunity against measles (OR 5.57; 95% CI 1.33-23.27; *p* 0.01), mumps (OR 5.03; 95% CI 1.66-15.23; *p* 0.004), VZV (OR 3.48; 95% CI 1.10-10.96; *p* 0.03), and tetanus (OR 7.02; 95% CI 1.66-29.8; *p* 0.008).

## Discussion

The findings of our study reveal intriguing differences in the persistence of humoral immune responses against exanthematous viruses and bacterial pathogens in two cohorts of adult-onset primary and secondary antibody deficiency.

First, our results suggest that IgG against exanthematous viruses, including measles, mumps, and varicella as well as tetanus, are preferentially retained compared to post-vaccination anti-pneumococcal and anti-Hib antibodies, both in CVID and SeAD patients. This discrepancy suggests that the factors influencing the maintenance of humoral immunity may vary between exanthematous live viral exposure and bacterial polysaccharide antigen exposure by immunisation or natural exposure. The observed differences in antibody persistence have potential implications for infection susceptibility and overall clinical outcomes of adult-onset antibody deficiency patients. The mechanistic explanation for this observation is uncertain but indicates significant differences in the physiology of anti-bacterial polysaccharide antibody memory responses versus immunological memory generated by live viral vaccines or natural infection, by exanthematous viruses, or repeated immunisation with an adjuvant supported bacterial toxoid. Investigating the mechanistic basis of this is likely to be fruitful and advance the development of achieving enduring protection induced by anti-viral vaccines.

It is widely acknowledged that bacterial capsular polysaccharides (CPS) are T cell-independent (TI) antigens, which stimulate specific B cells in the splenic marginal zone to produce antibodies capable of inducing opsonisation and consequent phagocyte mediated clearance (reviewed in (28) and (29)). In rodent studies, such immune responses do not induce significant responses of follicular B cells, or a germinal centre response and result in the generation of germline antibodies without mutation of V regions. In contrast, human splenic marginal zone B cells possess somatically mutated surface immunoglobulin receptors with high affinity to the cognate antigen. TI responses do not generate B cell memory and the responding B cells demonstrate isotype switching and transform into plasma cells secreting specific antibodies at a high rate, with a variable duration of in-vivo survival. Human infants below 2 years of age are physiologically incapable of responding to TI antigens.

Polysaccharide-protein conjugate (C-TI) vaccines, which overcome this physiological deficit, in-contrast, induce a T cell-dependent memory response, generated via a germinal centre response characterised by a switch to high-affinity IgG antibodies in infants from birth (30, 31). Neither the plain capsular polysaccharide nor C-TI vaccines, induce enduring memory antibody responses in older infants and adults(28, 31).

Thymus-dependent antigens are proteins which are processed within endosomes of antigen presenting cells to peptides to enable presentation to CD4+ T cells, when exhibited on MHC II molecules, thus initiating T cell activation essential for providing help to B cells responding to their cognate antigen (reviewed in (29)and (30)). In producing these responses follicular B cells become activated via signals like CD40-CD40 ligand interactions and cytokines IL-21 and IL-4 and guided by BCL6, form germinal centres, where isotype switching, affinity maturation and B memory cell generation occur. When follicular B cells have undergone affinity maturation and class-switching, they commit to secreted antibody production and leave secondary lymphoid tissues and home to the bone marrow. In this niche they terminally differentiate into plasma cells, which generate high-affinity, isotype-switched antibodies at a high rate. A fraction of affinity-maturated, class-switched follicular B cells exit germinal centres and become recirculating B memory cells. On re-exposure to cognate antigens as happens during community exposure to exanthematous viruses, memory B cells recognising cognate antigens can rapidly differentiate into plasma cells or re-initiate a new germinal centre response where they may undergo further affinity maturation.

It is now generally accepted that plasma cells can become long-lived (memory) plasma cells and secrete antibodies for months, years or a lifetime(32, 33).

It is striking that in CVID patients and most of those studied with SeAD, there’s a failure of response to the bacterial polysaccharide vaccine, pneumovax, and that haemophilus B C-TI, vaccine, although a T cell dependent vaccine, failed to induce an IgG Hib capsular polysaccharide specific antibody response. Therefore, the mechanisms of T cell priming, B memory cell generation, and enduring plasma cells seeding to the bone marrow, must be different from those initiated by, natural exanthematous viral infection, or attenuated live viral vaccines. With C-TI vaccines, the induction of the antibody response, is rate limited by the degree of pre-existent T cell priming. Since the CVID and SeAD, patients retain their IgG antibody response to tetanus toxoid, this cannot be the rate limiting factor for Hib capsular polysaccharide specific IgG antibody production, in patients reported in the study.

In this context, it is worth noting that with respect to the measles virus, the clearance of viral RNA from blood and tissues is much slower than clearance of infectious virus and proceeds over weeks to months after resolution of the rash(34). This period of RNA persistence may be responsible for priming for life-long protection from MeV re-infection, following natural MeV infection.

Following completion of immunisation with tetanus toxoid, which is formulated with adjuvant, according to recommended schedules, protective levels are very long lasting, implying the generation of long-lived plasma cell, as well as immunological memory(35). We have shown that this response is retained, despite the development of CVID, or SeAD, in adults.

One further consideration to add to the above is that despite anti-MMR IgG being apparently better conserved that anti-pneumococcal/anti-Hib IgG, caution should be employed when concluding that, positive antibody levels will be reflected by real-life significantly higher clinical immunity against these viral infections. This is supported from evidence of granulomatous disease being associated with these viruses in immunodeficient patients as already discussed, suggesting inadequate protection despite seropositivity. Furthermore, from a more technical point of view, the method used for IgG concentration quantification may limit such conclusions from being extrapolated. Specifically, in the case of anti-measles IgG, its measurement is widely done via ELISA given its technical convenience and standardisation (36). However, this method employs either the recombinant nucleoprotein or inactivated whole virus and hence may not detect antibodies recognising the surface glycoproteins H and F that are responsible for receptor attachment and host cell entry of the measles virus and are hence associated with clinical protection. Additionally, the benchmark concertation assumed to correlate with clinical measles immunity has been questioned. This baseline concentration is based on the evaluation of antibody titres that appeared to provide measles protection during an outbreak in a US educational institution (37). The antibody levels were measured by an ELISA method, which was standardised against the WHO measles antibody international standard, (currently the WHO 3rd international standard; NIBSC 97/648) and was determined to be ≥120 milli IU/ml. The WHO standard serum had been calibrated by comparing to a measles virus plaque neutralisation assay (PRNT) (38). A large systematic analysis highlighted the weakness of the assumptions that have assigned this concentration of antibody as equivalent to that capable of producing biological protection, to individuals in the community (39). In addition to this, the documentation accompanying the WHO reference standard, states : “this preparation has not been calibrated for use in ELISA assays and/or a unitage assigned for this use.”

The above illustrate that despite discovering an association regarding conservation or lack of these antibodies, translating this into the clinical context is challenging and would require further research to redefine at which concentration such protection is afforded, by determining concentrations capable of live virus neutralisation

A second observation of our study was the higher likelihood for retained immunity against exanthematous viruses in the SeAD cohort compared to CVID patients, even after adjusting for age and sex. This finding suggests that the cause of antibody deficiency may play a crucial role in shaping the immune response and raises questions about the impact of rituximab treatment on humoral immunity. Rituximab, a monoclonal antibody targeting CD20-positive B cells, is widely used in the treatment of autoimmune conditions. Morbidity and mortality due to vaccine-preventable illnesses, particularly infection by S. pneumoniae and H. influenzae, is higher among patients with autoimmune conditions compared with the general population (40), and vaccination against infections prior to disease modifying therapy, is recommended by most major national and international rheumatological societies. Our findings hint at the possibility that rituximab-treated individuals may retain better humoral responses against certain viral pathogens compared to patients with primary antibody immunodeficiency.

As measles resurfaces in developed countries (7), and given that patients with immunodeficiency are at risk for complications following both infection and vaccination with the live MMR vaccine (41), understanding the details of the dynamics of post-rituximab humoral immunity are crucial in shaping management strategies. Indications that IgG levels correlate with neutralising antibody activity (42), can offer reassurance that the presence of laboratory immunity to measles correlates with clinical protection.

Our study has certain limitations, including its relatively small cohort size, the assumption that all subjects were vaccinated against MMR in childhood, although this is generally universal in the UK population, and the observational nature of our data within a single centre. Further research, especially longitudinal studies with larger sample sizes, is needed to confirm and expand upon our findings. Additionally, investigations into the mechanisms underlying the observed differences in antibody persistence, especially in rituximab-treated individuals, are crucial for advancing our understanding of immune responses in adult-onset antibody deficiency.

In conclusion, our study sheds light on the differential maintenance of humoral immune responses in adult-onset antibody deficiency. The preferential persistence of immunity against common exanthematous viruses compared to anti-bacterial antibodies, particularly in rituximab-treated individuals, prompts further exploration of the underlying mechanisms and clinical implications. This knowledge is crucial for optimizing vaccination strategies and managing the risk of infectious complications in individuals with antibody deficiencies, especially in the context of immunomodulatory therapies.

## Materials and Methods

This was a cohort study looking at the seroprevalence of anti-exanthematous and anti-bacterial IgG in two cohorts of patients with common variable immunodeficiency (CVID) or secondary antibody deficiency (SeAD) within the remit of the Immunology Department at the Cambridge University Hospital, United Kingdom (UK), a tertiary department covering a large area in the East of England (approximate denominator population of 5 million).

### Data Collection

Anonymized baseline data of consecutively referred patients with suspected CVID or SeAD who had their diagnosis subsequently confirmed under the care of the Immunology Department at the Cambridge University Hospital, UK were gathered from medical records as well as from databases populated with pre-collected data stored within the Immunology department intranet. The types of data collected for each patient and the normal ranges for each value can be seen in Table II below. This anonymised retrospective service review was approved by Cambridge University Hospital NHS trust, Cambridge UK, Audit department (Reference: Clinical Project ID6338, 07/10/2024). In accordance with the UK National Health Service Research Ethics Committee guidelines, ethics approval was not required as this work comprises anonymous audit of retrospective data.

**Table II.** Type of data collected.

| Type of data | Range |
| --- | --- |
| Sex | Binary value (Male/Female) |
| Cause of immunodeficiency (if known) | Nominal value |
| Age at diagnosis | >18 years |
| IgG Level | Normal: 6.00 - 16.00 g/L<br>Undetectable if <1.4 g/L |
| IgM Level | Normal: 0.50 - 2.00 g/L<br>Undetectable if <0.2 g/L |
| IgA Level | Normal: 0.80 – 4.00 g/L<br>Undetectable <0.3 g/L |
| Pre-vaccination anti-pneumococcal IgG | Poor response to pneumococcal vaccine defined as IgG against <7/13 serovars postvaccination |
| Postvaccination anti-pneumococcal IgG |  |
| Pre-vaccination anti-Hib IgG | Poor response to Hib vaccine defined as <1.000 µg/ml IgG titre postvaccination |
| Postvaccination anti-Hib IgG |  |
| Anti-measles IgG | Qualitative value – Detected or Undetectable |
| Anti-mumps IgG | Qualitative value – Detected or Undetectable |
| Anti-rubella IgG | Qualitative value – Detected or Undetectable |
| Anti-varicella IgG | Adequate immunity if >165 mIU/ml<br>Undetectable if <10 mIU/ml |
| Anti-tetanus IgG | Adequate immunity if >0.15<br>Undetectable if <0.005 |

Hib: *Haemophilus Influenzae B*

### Vaccinations

During clinical evaluation to diagnose clinically significant antibody deficiency, all patients were assessed at baseline for pneumococcal and anti-*Haemophilus influenzae* type b (Hib) IgG and were subsequently vaccinated against these organisms via a single dose of a pneumococcal polysaccharide vaccine (Pneumovax23®), and a conjugated Hib and *Neisseria meningitidis* group C (Hib/MenC) vaccine (Menitorix®). The anti-bacterial IgG titres were measured again more than 6 weeks post vaccination to assess the patients’ immunological responses to vaccination. This approach is the accepted standard of care in evaluating patients for antibody immunodeficiency(25-27, 43-45).

### Data management and statistics

Data collection and management were carried out using Microsoft Excel. Chi-square tests were used as appropriate to explore whether seroprevalence was statistically different in distinct patient groups. Stata/SE 17.0 (Stata corporation) was used for t-test, univariate and multivariate analysis.

### Patient population

We collected serological and demographic data on 181 patients with clinically suspected CVID and 50 with suspected SeAD under the care of the Immunology Department at Addenbrooke’s Hospital. These were then validated to confirm the diagnosis in each case (Table III and Table IV). CVID diagnosis was confirmed using the European Society for Immunodeficiencies criteria for probable CVID diagnosis. After patients with incomplete records were excluded and all patients with diagnoses other than CVID were also eliminated (e.g. specific antibody deficiency), a cohort of 50 CVID and 49 SeAD patients was established.

**Table III.**
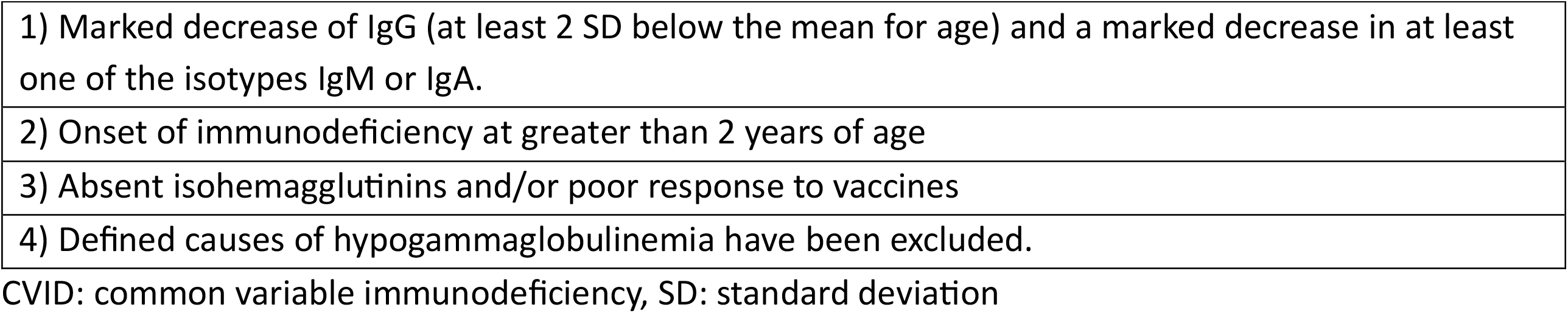
Validation Criteria for CVID Diagnosis.

|  |
| --- |
| 1) Marked decrease of IgG (at least 2 SD below the mean for age) and a marked decrease in at least one of the isotypes IgM or IgA. |
| 2) Onset of immunodeficiency at greater than 2 years of age |
| 3) Absent isohemagglutinins and/or poor response to vaccines |
| 4) Defined causes of hypogammaglobulinemia have been excluded. |

CVID: common variable immunodeficiency, SD: standard deviation

**Table IV.** Validation Criteria for SeAD Diagnosis.

|  |
| --- |
| 1) Marked decrease of IgG (at least 2 SD below the mean for age) and a marked decrease in at least one of the isotypes IgM or IgA. |
| 2) Onset of immunodeficiency at greater than 18 years of age |
| 3) Known secondary cause of hypogammaglobulinemia. |

SD: standard deviation, SeAD: secondary antibody deficiency

### Measurements

Immunoglobulin levels were measured at the Clinical Biochemistry and Immunology department at the Cambridge University Hospital, UK, (UK Accreditation Service, UKAS accredited).Specific antibody measurements against exanthematous viruses were performed at the Public Health England Clinical Microbiology and Public Health Laboratory, Cambridge, UK,(UK Accreditation Service, UKAS accredited.), using methods described below:

Serotype-specific pneumococcal antibody titres were measured to 13 of serotypes contained in Pneumovax II using a multiplex fluorescent bead assay (BioPlex Protein Array System, BioRad Laboratories, Hercules, CA), as described by Willcocks et al. (46). The Food and Drug Administration (FDA) standard reference serum (89SF) was used as calibrant in validating the serotype-specific anti-pneumococcal antibody levels. An adequate response to immunization, in adults was defined as having serotype specific antibody value ≥ 0.35 μg/mL (the consensus level considered to be protective against invasive pneumococcal disease in accordance with WHO consensus(47), to at least 7/13 serotypes tested which threshold had been determined to have (90% sensitivity, 88% specificity) for diagnosing antibody deficiency (unpublished data). We have also shown the serotype specific antibody responses evaluated against the US consensus criteria for determining an adequate adult response as a post-immunization titre >1.3 ug/ml (25).

Antibodies to Haemophilus influenzae type B (Hib) capsular polysaccharide and to tetanus toxoid were also measured using a multiplex fluorescent bead assay with the beads being conjugated to the Hib polysaccharide or tetanus toxoid respectively. These assays were calibrated against reference standards obtained from the institute of biological standards, UK. The optimum protective levels against Haemophilus B invasive disease considered to be 1 mcg/mL (48), and the minimum protective antibodies against tetanus considered to be 0.15 IU/mL (49).

The measles mumps, rubella and varicella antibodies were measured by the diagnostic virology laboratory at Cambridge University Hospital. Measles, mumps and varicella antibodies were measured using the Diasorin Liaison XL method (50) while rubella antibodies were measured using the Advia

### Centaur Rubella IgG assay

For the measles antibody assay results were generated relative to a calibrator curve on a scale of 5-300 Arbitrary Units (AU/mL) and the cut-off was 15.0 AU/mL. The cut-off value equates to 175 milli IU/mL using the WHO Third International Standard for anti-measles, NIBSC code: 97/648. (This data was supplied by the manufacturer of the assay). The measles plaque neutralisation titre (PNT) that corresponded to the protective titre of the WHO standard (97/648) was found to be ≥120 milli IU/ml in a study that reviewed protective immunity of college students after a measles outbreak (37) .However, the documentation of the standard NIBSC code: 97/648, states, “this preparation has not been calibrated for use in ELISA assays and/or a unitage assigned for this use.”. Patients that were labelled as measles antibody “not detected” in our study had values <5.0 AU/mL and patients that were labelled as ‘measles antibody detected’ were >300.00 AU/mL.

The LIAISON VZV IgG HT immunoassay (51) was used for measurement of anti-VZV IgG. In immunocompetent patients a result below 100 mIU/mL indicated absence of antibodies to VZV while a result above or equal to 100 mIU/mL was positive for the presence of these antibodies. The threshold for determining the positive threshold was higher in immunocompromised individuals with results above 165mIU/ml being reported as positive for evidence of VZV immunity. These criteria have been externally validated (52). The analytical sensitivity of this assay was determined using a series of serial dilutions of the WHO First International Standard for varicella zoster immunoglobulin (1987), NIBSC code: W1044 in negative serum matrix.

The LIAISON Mumps IgG immunoassay (53) was used for measurement of anti-mumps IgG. The assay range was 5 to 300 AU/mL mumps virus IgG. The cut-off value discriminating between the presence and the absence of mumps virus IgG was set as 11 AU/mL, with IgG concentrations below 9.0 AU/mL and those ranging between 9.0 and 11.0 AU/mL, which were graded indeterminate, being reported as negative (54).

For anti-rubella IgG, the Advia ***Centaur Rubella IgG assay*** (https://doclib.siemens-healthineers.com/rest/v1/view?document-id=637505) was used for measurement.. Expected values were established using the ADVIA Centaur system and confirmed by assay comparison with the UK National External Quality Assurance Scheme distribution for Rubella as external quality assurance method. A value of <10 IU/mL was reported as not detected while >10 IU/mL was reported as detected (55). These commercial immunoassays have been calibrated with a WHO international standard and report results in international units per millilitre (Anti-rubella Immunoglobulin, Human WHO International Standard RUBI-1-94).

## Data Availability

All data produced in the present study are available upon reasonable request to the authors.

## Abbreviations

CAR: chimeric antigen receptor
CLL: chronic lymphocytic leukemia
CVID: common variable immunodeficiency
Hib: *Haemophilus influenzae* type b
HIV: human immunodeficiency virus
IgG: immunoglobulin G
MGUS: monoclonal gammopathy of undetermined significance
MMR: measles-mumps-rubella virus
SeAD: secondary antibody deficiency
VZV: varicella zoster virus
ELISA: enzyme-linked immunosorbent assays

## References

1. Yung CF, Andrews N, Bukasa A, Brown KE, Ramsay M. Mumps complications and effects of mumps vaccination, England and Wales, 2002-2006. Emerg Infect Dis. 2011;17(4):661–7; quiz 766.

2. Bester JC. Measles and Measles Vaccination: A Review. JAMA Pediatr. 2016;170(12):1209–15.

3. Di Pietrantonj C, Rivetti A, Marchione P, Debalini MG, Demicheli V. Vaccines for measles, mumps, rubella, and varicella in children. Cochrane Database Syst Rev. 2021;11(11):CD004407.

4. Gershon AA, Gershon MD, Shapiro ED. Live Attenuated Varicella Vaccine: Prevention of Varicella and of Zoster. J Infect Dis. 2021;224(12 Suppl 2):S387–S97.

5. Sbarra AN, Mosser JF, Jit M, Ferrari M, Ramshaw RE, O’Connor P, et al. Estimating national-level measles case-fatality ratios in low-income and middle-income countries: an updated systematic review and modelling study. Lancet Glob Health. 2023;11(4):e516–e24.

6. Fragkou PC, Thomas K, Sympardi S, Liatsos GD, Pirounaki M, Sambatakou H, et al. Clinical characteristics and outcomes of measles outbreak in adults: A multicenter retrospective observational study of 93 hospitalized adults in Greece. J Clin Virol. 2020;131:104608.

7. Mathis AD, Raines K, Masters NB, Filardo TD, Kim G, Crooke SN, et al. Measles - United States, January 1, 2020-March 28, 2024. MMWR Morb Mortal Wkly Rep. 2024;73(14):295–300.

8. Laing KJ, Ouwendijk WJD, Koelle DM, Verjans G. Immunobiology of Varicella-Zoster Virus Infection. J Infect Dis. 2018;218(suppl_2):S68–S74.

9. Naniche D. Human immunology of measles virus infection. Curr Top Microbiol Immunol. 2009;330:151–71.

10. Thompson KM, Odahowski CL. Systematic Review of Measles and Rubella Serology Studies. Risk Anal. 2016;36(7):1459–86.

11. Pauksen K, Duraj V, Ljungman P, Sjolin J, Oberg G, Lonnerholm G, et al. Immunity to and immunization against measles, rubella and mumps in patients after autologous bone marrow transplantation. Bone Marrow Transplant. 1992;9(6):427–32.

12. Kawamura K, Wada H, Nakasone H, Akahoshi Y, Kawamura S, Takeshita J, et al. Immunity and Vaccination Against Measles, Mumps, and Rubella in Adult Allogeneic Hematopoietic Stem Cell Transplant Recipients. Transplant Cell Ther. 2021;27(5):436 e1–e8.

13. Rezahosseini O, Sorensen SS, Perch M, Ekenberg C, Moller DL, Knudsen AD, et al. Measles, Mumps, Rubella, and Varicella Zoster Virus Serology and Infections in Solid Organ Transplant Recipients During the First Year Posttransplantation. Clin Infect Dis. 2021;73(11):e3733–e9.

14. Hill JA, Krantz EM, Hay KA, Dasgupta S, Stevens-Ayers T, Bender Ignacio RA, et al. Durable preservation of antiviral antibodies after CD19-directed chimeric antigen receptor T-cell immunotherapy. Blood Adv. 2019;3(22):3590–601.

15. Loevinsohn G, Rosman L, Moss WJ. Measles Seroprevalence and Vaccine Responses in Human Immunodeficiency Virus-infected Adolescents and Adults: A Systematic Review. Clin Infect Dis. 2019;69(5):836–44.

16. Kuijpers TW, Weening RS, Out TA. IgG subclass deficiencies and recurrent pyogenic infections, unresponsiveness against bacterial polysaccharide antigens. Allergol Immunopathol (Madr). 1992;20(1):28–34.

17. Rijkers GT, Sanders LA, Zegers BJ. Anti-capsular polysaccharide antibody deficiency states. Immunodeficiency. 1993;5(1):1–21.

18. Notarangelo LD. Rubella Virus-Associated Granulomas in Immunocompetent Adults-Possible Implications. JAMA Dermatol. 2022;158(6):611–3.

19. Perelygina L, Icenogle J, Sullivan KE. Rubella virus-associated chronic inflammation in primary immunodeficiency diseases. Curr Opin Allergy Clin Immunol. 2020;20(6):574–81.

20. Pei S, Khazaeli M, Hao L, Chen MH, Perelygina L, Kuraitis D. Rubella virus-associated necrotizing neutrophilic granuloma in a patient with common variable immunodeficiency. J Cutan Pathol. 2023;50(11):971–6.

21. Bender NR, Cardwell LA, Siegel D, Sokumbi O. Rubella Vaccine Persistence Within Cutaneous Granulomas in Common Variable Immunodeficiency Disorder. Am J Dermatopathol. 2020;42(6):455–7.

22. ten Berge JC, van Daele PL, Rothova A. Rubella Virus-associated Anterior Uveitis in a Vaccinated Patient: A Case Report. Ocul Immunol Inflamm. 2016;24(1):113–4.

23. https://esid.org/working-parties/registry-working-party/diagnosis-criteria/. ESID Registry - Working definitions for clinical diagnosis of PID Accessed 03/03/2025 [

24. Browning MJ, Lim MT, Kenia P, Whittle M, Doffinger R, Barcenas-Morales G, et al. Pneumococcal polysaccharide vaccine responses are impaired in a subgroup of children with cystic fibrosis. J Cyst Fibros. 2014;13(6):632–8.

25. Orange JS, Ballow M, Stiehm ER, Ballas ZK, Chinen J, De La Morena M, et al. Use and interpretation of diagnostic vaccination in primary immunodeficiency: a working group report of the Basic and Clinical Immunology Interest Section of the American Academy of Allergy, Asthma & Immunology. J Allergy Clin Immunol. 2012;130(3 Suppl):S1–24.

26. Hansen AT, Söderström A, Jørgensen CS, Larsen CS, Petersen MS, Bernth Jensen JM. Diagnostic Vaccination in Clinical Practice. Frontiers in Immunology. 2021;12.

27. Martins TB, Hill HR, Peterson LK. Evaluating patient immunocompetence through antibody response to pneumococcal polysaccharide vaccine using a newly developed 23 serotype multiplexed assay. Clin Immunol. 2024;265:110295.

28. Weller S, Reynaud C-A, Weill J-C. Vaccination against encapsulated bacteria in humans: paradoxes. Trends in Immunology. 2005;26(2):85–9.

29. https://www.uptodate.com/contents/the-adaptive-humoral-immune-response?csi=d08f11dd-54bd-407c-a505-ca8389e0040d&source=contentShare#H2399465225. UpToDate - The Adaptive humoral immune response Accessed 11/03/2025 [

30. Pollard AJ, Bijker EM. A guide to vaccinology: from basic principles to new developments. Nature Reviews Immunology. 2021;21(2):83–100.

31. Pollard AJ, Perrett KP, Beverley PC. Maintaining protection against invasive bacteria with protein–polysaccharide conjugate vaccines. Nature Reviews Immunology. 2009;9(3):213–20.

32. Hammarlund E, Thomas A, Amanna IJ, Holden LA, Slayden OD, Park B, et al. Plasma cell survival in the absence of B cell memory. Nat Commun. 2017;8(1):1781.

33. Khodadadi L, Cheng Q, Radbruch A, Hiepe F. The Maintenance of Memory Plasma Cells. Front Immunol. 2019;10:721.

34. Griffin DE. The Immune Response in Measles: Virus Control, Clearance and Protective Immunity. Viruses. 2016;8(10).

35. Pool V, Tomovici A, Johnson DR, Greenberg DP, Decker MD. Humoral immunity 10 years after booster immunization with an adolescent and adult formulation combined tetanus, diphtheria, and 5-component acellular pertussis vaccine in the USA. Vaccine. 2018;36(17):2282–7.

36. Lutz CS, Hasan AZ, Bolotin S, Crowcroft NS, Cutts FT, Joh E, et al. Comparison of measles IgG enzyme immunoassays (EIA) versus plaque reduction neutralization test (PRNT) for measuring measles serostatus: a systematic review of head-to-head analyses of measles IgG EIA and PRNT. BMC Infect Dis. 2023;23(1):367.

37. Chen RT, Markowitz LE, Albrecht P, Stewart JA, Mofenson LM, Preblud SR, et al. Measles Antibody: Reevaluation of Protective Titers. The Journal of Infectious Diseases. 1990;162(5):1036–42.

38. Cohen BJ, Doblas D, Andrews N. Comparison of plaque reduction neutralisation test (PRNT) and measles virus-specific IgG ELISA for assessing immunogenicity of measles vaccination. Vaccine. 2008;26(50):6392–7.

39. Bolotin S, Hughes SL, Gul N, Khan S, Rota PA, Severini A, et al. What Is the Evidence to Support a Correlate of Protection for Measles? A Systematic Review. The Journal of Infectious Diseases. 2019;221(10):1576–83.

40. Wakabayashi A, Ishiguro T, Takaku Y, Miyahara Y, Kagiyama N, Takayanagi N. Clinical characteristics and prognostic factors of pneumonia in patients with and without rheumatoid arthritis. PLoS One. 2018;13(8):e0201799.

41. Rubin LG, Levin MJ, Ljungman P, Davies EG, Avery R, Tomblyn M, et al. 2013 IDSA clinical practice guideline for vaccination of the immunocompromised host. Clin Infect Dis. 2014;58(3):309–18.

42. Hu S, Logan N, Coleman S, Evans C, Willett BJ, Hosie MJ. Correlating IgG Levels with Neutralising Antibody Levels to Indicate Clinical Protection in Healthcare Workers at Risk during a Measles Outbreak. Viruses. 2022;14(8).

43. Chapel HM, Consensus Panel For The D, Management Of Primary Antibody D. Consensus On Diagnosis And Management Of Primary Antibody Deficiencies. BMJ: British Medical Journal. 1994;308(6928):581–5.

44. Grigoriadou S, Clubbe R, Garcez T, Huissoon A, Grosse-Kreul D, Jolles S, et al. British Society for Immunology and United Kingdom Primary Immunodeficiency Network (UKPIN) consensus guideline for the management of immunoglobulin replacement therapy. Clinical and Experimental Immunology. 2022;210(1):1–13.

45. Jolles S, Michallet M, Agostini C, Albert MH, Edgar D, Ria R, et al. Treating secondary antibody deficiency in patients with haematological malignancy: European expert consensus. Eur J Haematol. 2021;106(4):439–49.

46. Willcocks LC, Chaudhry AN, Smith JC, Ojha S, Doffinger R, Watson CJ, et al. The effect of sirolimus therapy on vaccine responses in transplant recipients. Am J Transplant. 2007;7(8):2006–11.

47. Siber GR, Chang I, Baker S, Fernsten P, O’Brien KL, Santosham M, et al. Estimating the protective concentration of anti-pneumococcal capsular polysaccharide antibodies. Vaccine. 2007;25(19):3816–26.

48. Käyhty H, Peltola H, Karanko V, Mäkelä PH. The protective level of serum antibodies to the capsular polysaccharide of Haemophilus influenzae type b. J Infect Dis. 1983;147(6):1100.

49. Gergen PJ, McQuillan GM, Kiely M, Ezzati-Rice TM, Sutter RW, Virella G. A Population-Based Serologic Survey of Immunity to Tetanus in the United States. New England Journal of Medicine. 1995;332(12):761–7.

50. https://int.diasorin.com/en/immunodiagnostics/tools/liaison-xl?f%5B0%5D=sapl%3A44. LIAISON XL. Accessed 03/03/2025.

51. https://www.accessdata.fda.gov/cdrh_docs/reviews/K231214.pdf. LIAISON VZV IgG HT. Accessed 03/03/2025.

52. Maple PA, Haedicke J, Quinlivan M, Steinberg SP, Gershon AA, Brown KE, et al. The differences in short- and long-term varicella-zoster virus (VZV) immunoglobulin G levels following varicella vaccination of healthcare workers measured by VZV fluorescent-antibody-to-membrane-antigen assay (FAMA), VZV time-resolved fluorescence immunoassay and a VZV purified glycoprotein enzyme immunoassay. Epidemiol Infect. 2016;144(11):2345–53.

53. https://int.diasorin.com/en/immunodiagnostics/infectious-diseases/mumps-virus. LIAISON® Mumps Diagnostic Solution. Accessed 03/03/2025.

54. Ferrari C, Trabucco Aurilio M, Mazza A, Pietroiusti A, Magrini A, Balbi O, et al. Evaluation of Immunity for Mumps among Vaccinated Medical Students. Vaccines (Basel). 2021;9(6).

55. Bouthry E, Furione M, Huzly D, Ogee-Nwankwo A, Hao L, Adebayo A, et al. Assessing Immunity to Rubella Virus: a Plea for Standardization of IgG (Immuno)assays. J Clin Microbiol. 2016;54(7):1720–5.

